# Multilevel Determinants of Teenage Pregnancies Among Girls Aged 13-19 Years in Rural Uganda: Community Based Cross Sectonal Study

**DOI:** 10.1101/2025.09.30.25337036

**Authors:** Wandeka Joyce Wamakote, Ronald Kooko, Simon P.S Kibira, Justine Bukenya, Rogers Kisame

**Affiliations:** Department of Community Health and Behavioral Science, School of public health, Makerere University, Kampala, Uganda; Ministry of Health-Uganda, Department of Integrated Epidemiology, Surveillance and Public Health Emergencies; Baylor College of Medicine Children’s Foundation Uganda, Kampala, Uganda

**Keywords:** Teenage pregnancy, early marriage, domestic violence, predictors

## Abstract

**Background:** Teenage pregnancy is a global public health problem with serious social and medical implications relating to maternal and child health. In Uganda, particularly Hoima district, limited information about the factors influencing teenage pregnancy is available. This study estimated the prevalence of teenage pregnancy and associated factors among teenage girls aged 13-19 years to guide design the interventions to mitigate this problem.

**Methods:** This was a community-based cross-sectional study that utilized multi-stage random sampling approach to select 543 study respondents. Data were collected using a pre-tested interviewer-administered structured questionnaire and analyzed using SPSS V26. Multivariable logistic regression analysis was conducted to determine independent predictors of teenage pregnancy among girls 13-19 years.

**Results:** The prevalence of teenage pregnancy was 29%. Age (18-19 years) (AOR 4.4, 95%CI 1.52–12.78), low parents’ economic status (AOR 5.4, 95%CI 2.47–11.78), multiple sexual partners (AOR 8.0, 95%CI 4.51–14.18), being out of school (AOR 12, 95%CI 5.02–29.06), early marriage (AOR 37, 95%CI 13.35–107.54), having no control over sex (AOR 47, 95%CI 13.7-163.8), not discussing SRH with parents (AOR 8.4, 95%CI 3.31–21.51), witnessed domestic violence (AOR 30, 95%CI 11.97–77.53), never received counselling (AOR 5.7, 95%CI 3.56–9.01), and, rural residence (AOR 1.8, 95%CI 1.15–2.94) were significant predictors of teenage pregnancy.

**Conclusion:** Teenage pregnancy is highly prevalent suggesting that teenage pregnancy is still a public health and social problem in Hoima district, Uganda. It is imperative for the Ministry of Health (MOH) and implementing partners to scale up and expand teenage friendly health services, consolidate efforts towards keeping girls in school, strengthen the policy of delaying early/child marriage, improve on the economic welfare of households through job creation and skill the youth for self-employment.

## Introduction

The United Nations International Children’s Emergency Fund (UNICEF), defines teenage pregnancy as a pregnancy in girls within the ages of 13–19 years [1]. Teenage pregnancy is a worldwide problem bearing serious social and medical implications relating to maternal and child health [2]. It is estimated that about 21 million girls 15–19 years old get pregnant in low– and middle-income countries (LMICs) each year, of which approximately 50% are unintended resulting in an estimated 12 million births [3].

The highest teenage pregnancy rates are recorded in Africa. The overall pooled prevalence of adolescent pregnancy in Africa, Sub-Saharan Africa, and East Africa is 18.8%, 19.3% and, 21.5% respectively [4]. Uganda, with an estimated 24% teenage pregnancy, it has one of the highest rates of teenage pregnancies in sub-Saharan Africa [5]. Teenage pregnancy thus remains a burden, to both the community and the government of Uganda especially in terms of expenditure in an attempt to curb the detrimental effects of teenage pregnancy on the lives of teenagers [6]. The Eastern and East Central regions are reported to have the highest rates of teenage pregnancy in Uganda with 30.1% and 31.6% respectively which is higher than its surrounding regions like Karamoja and West Nile [7].

Early marriage, early initiation of sex and lack of access to reproductive health information and services are the leading drivers of adolescent pregnancy in the world [1, 8]. Teenage pregnancies may occur as a result of other factors: customs and traditions, adolescent sexual behavior, which may also be influenced by alcohol and drugs, lack of education and information about reproductive sexual health, lack of access to tools that prevent pregnancies, peer pressure to engage in sexual activity, incorrect use of contraception, sexual abuse like rape, poverty, exposure to abuse, violence, family strife at home, low self-esteem and low educational ambitions or goals [3]. There is growing evidence of associations between teenage motherhood and poor health outcomes [9]. Although adolescent birthrates have decreased globally from 64.5 births per 1000 women (15–19 years) in 2000 to 41.3 births per 1000 women in 2023, Sub-Saharan Africa region continue to have rates globally at 99.4 births per 1000 women in 2022 [10].

Despite the implementation policies and related laws such as the Uganda’s National Adolescent Reproductive Health Policy (2004) emphasizing reduction of adolescent pregnancies by increasing access to reproductive health services, the Penal Code Act criminalizing sexual exploitation of minors, and the school health policy that advocates for comprehensive sex education to mitigate early pregnancies, teenage pregnancies remain relatively high in Uganda, especially in Hoima district [11, 12].

Therefore, to address the impact of teenage pregnancies, there is a need to continuously understand the risk factors associated with teenage pregnancy. Despite the high prevalence of teenage pregnancy in Uganda generally and Hoima district in western Uganda in particular, previous research on the sexual and reproductive health was limited to in-school teenage girls’ population with less focus on community-based studies hence neglecting the unfortunate teenagers who may be out of school for various reasons.

Even though numerous studies have been done worldwide to identify a wide variety of risk factors associated with teenage pregnancies, there is need for the same to be done in the local context and enable the formulation of feasible national strategies to curb teenage pregnancies. This study therefore assessed the prevalence of teenage pregnancies and associated factors among teenagers in Hoima district, Uganda. The findings from this study provided information that can be used by government institutions, health administrators and other relevant stakeholders to strengthen the implementation of the existing laws around national health policy, school health policy, national adolescent health policy and penal code act among others. Furthermore, it also provided policymakers with context-specific information for formulating policies that promote education and support sexual and reproductive rights of teenage girls in Hoima district and Uganda at large

## Materials and Methods

### Study design and setting

This was a community-based cross sectional study involving quantitative data collection approaches in three sub-counties of Hoima district: Buseruka, Buhanika, and Kitoba.

### Study population

Teenage girls aged 13–19 years who had resided in the selected areas for at least 12 months were eligible.

### Sample size and sampling

Using the Kish Leslie formula, with a prevalence of 30.6% from a previous study [12], a 5% margin of error, 10% non-response rate and a design effect of 1.5, the sample size was calculated at 543. Multi-stage random sampling was used to select study respondents. Hoima district is divided into two counties, Bugahya and Kigorobya. The counties are sub divided into 5 sub counties and 1 Town Council namely; Bugahya County – Buseruka, Buhanika, Kitoba and Kyabagambire sub counties; Kigorobya County – Kigorobya Sub County and Kigorobya Town Council. Three sub counties (Buseruka, Buhanika, Kitoba) were randomly selected by simple random sampling using rotary method to participate in the study. The principal investigator then randomly selected half of the parishes from each selected sub-county. Here the names of all parishes in each selected sub-county were written on individual pieces of paper, folded, and placed in a container. Half of the parishes were then drawn from the container without replacement for participation in the study and this sampling technique was also applied at parish level to select half the number of villages in each parish to participate in the study. From each village, a list of households was obtained and the households to participate in the study were purposively selected. The number of teenagers from each selected village to participate in the study was determined by probability proportionate to size (PPS).

## Study variables

The primary outcome was teenage pregnancy among girls aged 13–19 years, defined as being currently pregnant or having been pregnant in the past 12 months. Participants answering “yes” to either question were coded as 1 (pregnant) and “no” as 0 (not pregnant). Independent variables included socio-demographic, individual, community, family, and health facility factors.

## Data collection

Trained research assistants with support from the principal investigator, local chairpersons and village health teams collected data for a period of one month from 6^th^ March 2024 to 6^th^ April 2024 using a pre-tested interviewer administered structured questionnaire. The questionnaire was developed from literature reviewed [7, 12].

## Quality control measures

The principal investigator selected the research assistants, trained them on questionnaire administration, data collection, and ethical considerations; they were also trained in effective communication skills, showing empathy and never being judgmental. The research assistants were aged between 20 and 30 years and fluent in the local language. This helped to ensure free, open communication for the participants. The data collection tools were pre-tested in Kakumiro district because of its resemblance in socio-demographic characteristics to Hoima and translated into Runyoro for relevance and clarity. Any inconsistencies detected at the pretesting stage were corrected prior to the actual data collection. Fieldwork was overseen by the principal investigator to ensure completeness and accuracy of data collected. Besides, data collected was edited and cross-checked regularly to align with the study objectives.

## Data analysis

Data was analyzed using SPSS V26. Analysis was carried out at univariable, bivariable and multivariable levels. At univariable level, analysis was ran for all the variables. Categorical data was presented in form of frequencies and percentages. Mean, median and mode were used to present continuous data. At bivariate level logistic regression was be used to model the association between teenage pregnancy and different factors producing crude odds ratios and P-values at 95% confidence intervals. P-value of less than 0.05 was used as a cut off for statistical significance. At multivariable level, variables that showed significant associations with the dependent variable at bivariate analysis were included into the multivariate model (P<0.05). Hosmer-Lemeshow goodness fit test was used to evaluate the overall model fitness of the final regression model

## Ethical considerations

This study was reviewed and approved by the Makerere University School of Public Health Research and Ethics Committee. Permission to conduct the study was also obtained from Hoima Distict Health Officer (DHO) and the District Health Team (DHT). Before recruitment, the purpose and procedures of the study were explained to potential participants, and participation was voluntary. Written informed consent was obtained from all adult participants (≥18 years) and for participants younger than 18 years, written informed consent was obtained from their parents or legal guardians, and assent was obtained from the minors themselves prior to completing the questionnaire. Confidentiality was maintained throughout the study, and all information collected was used solely for research purposes.

## Results

### Sociodemographic characteristics of participants

In this study, data from 543 participants were analyzed. The mean age was 15.9 ± 1.9 years and the median age was 16 years, with an interquartile range of 14–17 years old.

The majority of participants (61.1%) included in this study were living in rural areas, over 44.0 % were between the ages of 15–17 years and 24.1% between the ages of 18–19 years, majority of the respondents 58.8% had attained primary level of education, 33.0 % of them were married and 39.0 % had multiple sexual partners.

About two-thirds, (77.2%) of the parents whose teens were included the study had low/poor economic status, a high number (66.1%) of participants had both parents with Catholic as the dominant religion at 34.4%. Approximately, (44.4) % of the teens had witnessed domestic violence. There was almost a uniform sample distribution among the sub-counties, Buhanika, Buseruka and Kitoba at 34.4%, 33.0% and 32.6% respectively (Table 1)

**Table 1.**
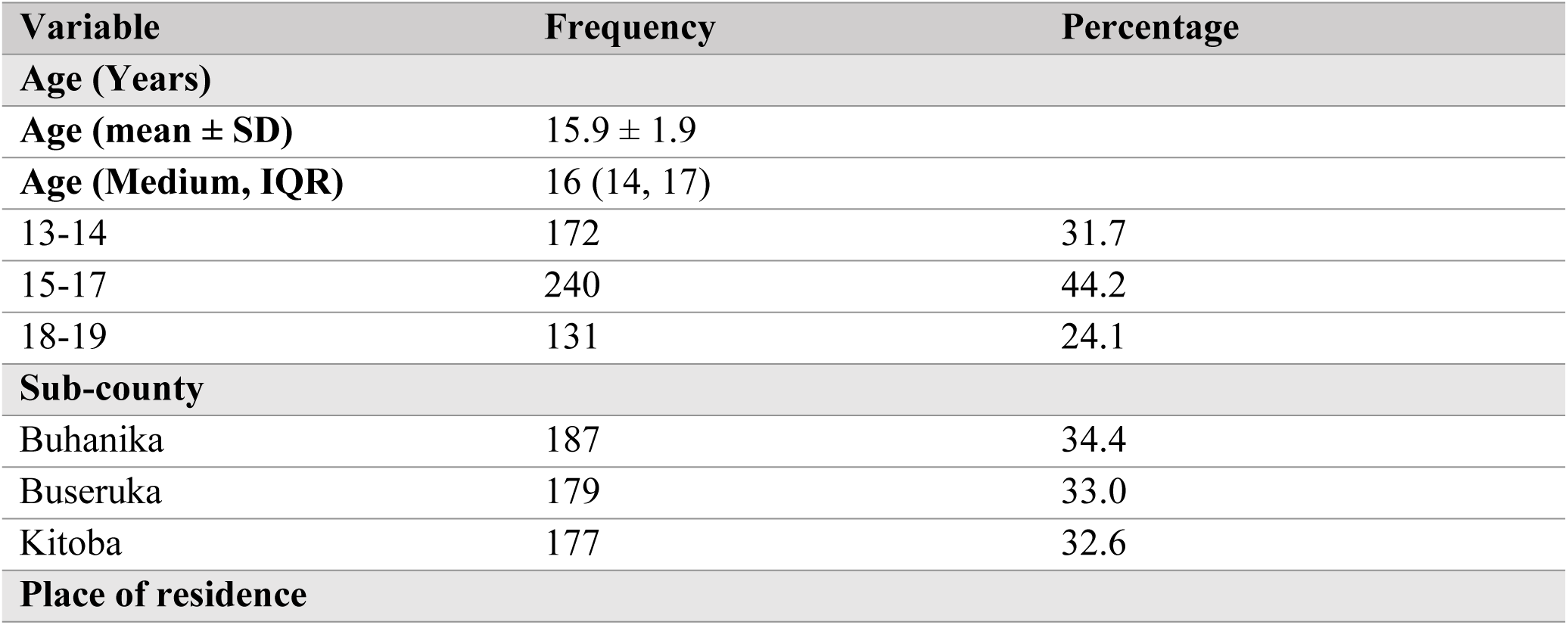
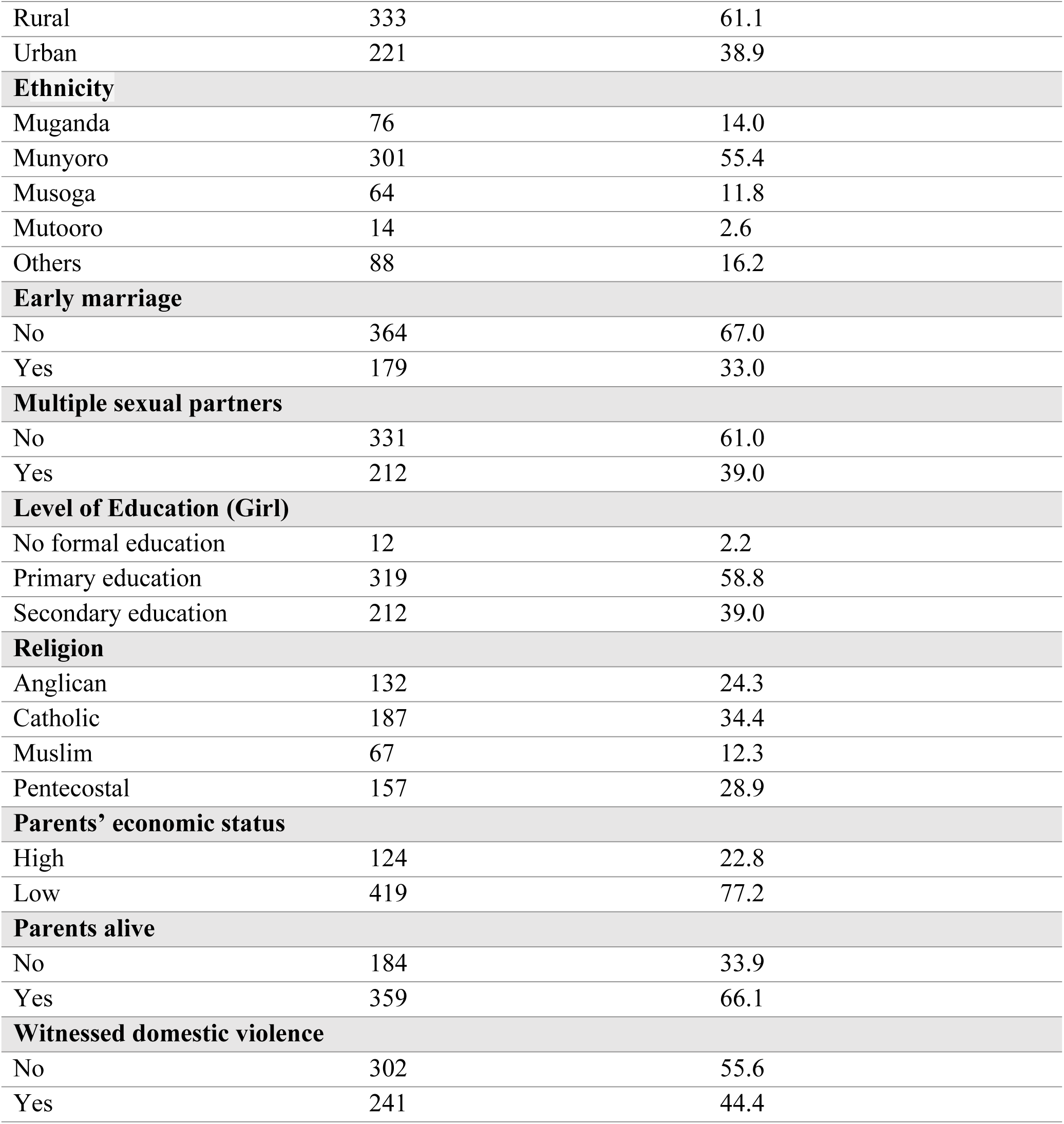
Socio-demographic characteristics of the respondents (N = 543)

### Prevalence of teenage pregnancies in Hoima district

Out of 543 teenagers, 158 (29%) had ever been pregnant (Figure 1).

**Figure 1.**
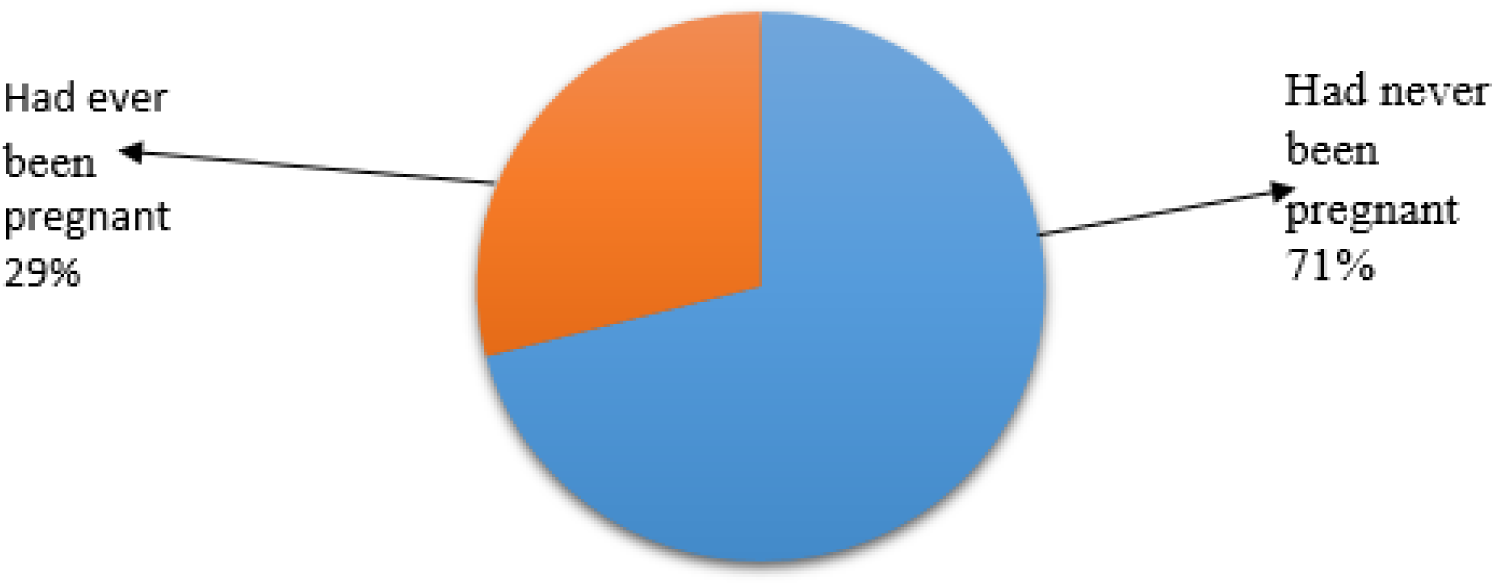
The prevalence of teenage pregnancies in the Hoima district, Uganda (N = 543)

### Factors associated with teenage pregnancies in Hoima district (Unadjusted analysis) Sociodemographic characteristics of respondents

Table 2. Presents the results of bivariate analysis of teenager and caretaker characteristics associated with teenage pregnancy. In this study, age was significantly associated with teenage pregnancy. There was a significantly high prevalence of teenage pregnancy among teens aged 18-19 at 64.9% compared to 4.7% among those aged 13-14. Teens aged 18-19 were 37 times more likely to get pregnant as compared to teens aged 13-14 (OR 37.0, 95%CI 17.11-83.90). The prevalence of teenage pregnancy among teenagers who were married was high at 74.1 % and 25 times more likely to get pregnant than their counterparts who were not married (OR 25.0, 95%CI 15.73-41.57). The education of the caretaker/parent was significantly associated with teenage pregnancy with about 45.6 % pregnancies happening among teenagers whose parents had no formal education and 18.1 % among those whose parents had at least attained secondary level of education.

**Table 2.** Unadjusted analysis of the sociodemographic characteristics associated with teenage pregnancy.

| Variable | Teenage pregnancy |  | Crude OR (95% CI) | P-value |
| --- | --- | --- | --- | --- |
|  | No N= 385 (%) | Yes N= 158 (%) |  |  |

| **Age groups (Years)** |  |  |  |  |
| --- | --- | --- | --- | --- |
| 13-14 | 164 (95.3) | 8 (4.7) | Reference |  |
| 15-17 | 175 (72.9) | 65 (27.1) | 7.6 (3.55 - 16.36 ) | < .001 |
| 18-19 | 46 (35.1) | 85 (64.9) | 37.9 (17.10 - 83.90) | < .001 |
| **Sub-county** |  |  |  |  |
| Buhanika | 126 (67.4) | 61 (32.6) | Reference |  |
| Buseruka | 133 (74.3) | 46 (25.7) | 0.7 (0.45 – 1.13) | 0.146 |
| Kitoba | 126 (71.2) | 51 (28.8) | 0.8 (0.54 – 1.31) | 0.432 |
| **Religion** |  |  |  |  |
| Anglican | 97 (73.5) | 35 (26.5) | Reference |  |
| Catholic | 134 (71.7) | 53 (28.3) | 1.1 (0.67 – 1.81) | 0.719 |
| Muslim | 43 (64.2) | 24 (35.8) | 1.5 (0.82 – 2 .91) | 0.176 |
| Pentecostal | 111 (70.7) | 46 (29.3) | 1.1 (0.67 - 1.93) | 0.600 |
| **Education level of Teenager** |  |  |  |  |
| No formal education | 8 (66.7) | 4 (33.3) | Reference |  |
| Primary education | 237 (74.3) | 82 (25.7) | 0.7 (0.20 – 2.34) | 0.556 |
| Secondary education | 140 (66.0) | 72 (34.0) | 1.0 (0.30 – 3.53) | 0.964 |
| **Marital status of Teenager** |  |  |  |  |
| Never married | 343 (90.0) | 38 (10.0) | Reference |  |
| Currently Married | 42 (25.9) | 120 (74.1) | 25 (15.73 – 41.57) | < .001 |
| **Marital status of Parents** |  |  |  |  |
| Currently married | 239 (68.5) | 110 (31.5) | Reference |  |
| Never married | 19 (95.0) | 1 (5.0) | 0.1 (0.02 – 0.87) | 0.036 |
| Separated | 127 (73.0) | 47 (27.0) | 0.8 (0.54 – 1.20) | 0.290 |
| **Education level of caretaker** |  |  |  |  |
| No formal education | 62 (54.4) | 52 (45.6) | Reference |  |
| Primary | 183 (70.9) | 75 (29.1) | 0.5 (0.31 – 0.77) | 0.002 |
| Secondary | 140 (81.9) | 31 (18.1) | 0.3 (0.15 – 0.45) | < .001 |
| **Biological parents alive** |  |  |  |  |
| Yes | 302 (84.1) | 57 (15.9) | Reference |  |
| No | 83 (45.1) | 101 (54.9) | 6.4 (4.30 – 9.67) | < .001 |
| **Parents' economic status** |  |  |  |  |
| High | 110 (88.7) | 14 (11.3) | Reference |  |
| Low | 275 (65.6) | 144 (34.4) | 4.1 (2.28 – 7.43) | < .001 |

\*significant at $P$ value $<0.05$ , Abbreviations: OR, odds ratio; CI, confidence interval.

From table three, there was a substantially high proportion of teenage pregnancies 34.4% among teenagers whose parents fall under a low (poor) economic status strata compared to 11.3% whose parents belong to a high (rich) economics status. Teenagers whose parents were poor were 4.1 times more likely to report having ever been pregnant compared to those whose parents were rich (OR 4.1, 95%CI 2.28-7.43). More than half of teenage pregnancies 54.9% were among teenagers who did not have both biological parents alive and these teenagers were 6.4 times more like to get pregnant than their counterparts who had both parents alive (OR 6.4, 95%CI 4.3-9.7) (Table 2).

Tables 3 present results of bivariate analysis of factors (individual, family, community and health facility) associated with teenage pregnancy.

### Individual factors

At a bivariate level, majority of the factors were significantly associated with teenage pregnancy except family planning use (p-value 0.149). There was a significantly high teenage pregnancy prevalence of 74.7% among teenagers with multiple sexual partners compared to 21.1% among teenagers with no multiple partners. Those with multiple sexual partners were 9.1 times more likely to get pregnant compared to their peers with only one sexual partner (OR 9.1, 95%CI 5.96-14.00). Similarly, teenagers who were not in school were 26 times more likely to get pregnant than their peers who were in school (OR 26, 95%CI 15.90-44.00)

Additionally, other individual factors that were associated with teenage pregnancy included early marriage, control over sex, sex for material support, family planning awareness (Table 3).

**Table 3.** Unadjusted analysis analysis of factors (Individual, Family, Community and Health facility) associated with teenage pregnancy N=543.

| Variable | Teenage pregnancy |  | Crude OR (95%CI) | P-value |
| --- | --- | --- | --- | --- |
|  | No N= 385 (%) | Yes N=158 (%) |  |  |
| Individual factors |  |  |  |  |
| Multiple sexual partners |  |  |  |  |
| No | 291 (87.9) | 40 (12.1) | Reference |  |
| Yes | 94 (25.3) | 118 (74.7) | 9.1 (5.96 – 14.00) | < .001 |
| School attendance |  |  |  |  |
| In school | 318 (93.0) | 24 (7.0) | Reference |  |
| Not in school | 67 (33.3) | 134 (66.7) | 26 (15.94 – 44.05) | < .001 |
| **Early marriage** |  |  |  |  |
| No | 341 (93.7) | 23 (6.3) | Reference |  |
| Yes | 44 (14.6) | 135 (85.4) | 45 (26.45 – 78.24) | < .001 |
| **Control over sex** |  |  |  |  |
| Yes | 379 (84.8) | 68 (15.2) | Reference |  |
| No | 6 (6.3) | 90 (93.8) | 83 (35.17 – 198.71) | < .001 |
| **Sex for material exchange** |  |  |  |  |
| No | 306 (86.7) | 47 (13.3) | Reference |  |
| Yes | 79 (41.6) | 111 (58.4) | 9 (6.0 – 13.0) | < .001 |
| **Family planning awareness** |  |  |  |  |
| Yes | 297 (80.5) | 72(19.5) | Reference |  |
| No | 88 (50.6) | 86 (49.4) | 4 (2.72 – 5.97) | < .001 |
| **Family planning use** |  |  |  |  |
| Yes | 233 (77.3) | 85 (26.7) | Reference |  |
| No | 152 (67.6) | 73 (32.4) | 1.3 (0.91 – 1.91) | 0.149 |
| **Family related factors** |  |  |  |  |
| **Discuss SRH with parents** |  |  |  |  |
| Yes | 273 (95.1) | 14 (4.9) | Reference |  |
| No | 112 (43.8) | 144 (56.3) | 25(13.88 – 45.28) | < .001 |
| **Parents talk about sex education** |  |  |  |  |
| Yes | 267 (95.0) | 14 (5.0) | Reference |  |
| No | 118 (45.0) | 144 (55.0) | 23 (12.90 – 41.98) | < .001 |
| **Witnessed domestic violence** |  |  |  |  |
| No | 293 (97.0) | 9 (3.0) | Reference |  |
| Yes | 92 (38.2) | 149 (61.8) | 52 (25.87 – 107.50) | < .001 |
| **Neglected by parents/caretaker** |  |  |  |  |
| No | 321 (83.8) | 62 (16.2) | Reference |  |
| Yes | 64 (40.0) | 96 (60.0) | 7.7 (5.12 – 11.79) | < .001 |
| **Stays with parents** |  |  |  |  |
| Yes | 324 (90.0) | 36 (10.0) | Reference |  |
| No | 61 (33.3) | 122 (66.7) | 18 (11.34 – 28.56) | < .001 |
| **Socioeconomic status of family** |  |  |  |  |
| High | 110 (88.7) | 14 (11.3) | Reference |  |
| Low | 275 (65.6) | 144 (34.4) | 4.1 (2.28 – 7.43) | < .001 |
| **Health facility factors** |  |  |  |  |
| **Provision of youth tailored health services** |  |  |  |  |
| Yes | 339 (76.0) | 107 (24.0) | Reference |  |
| No | 46 (47.4) | 51 (52.6) | 3.5 (2.23 – 5.53) | < .001 |
| **Affordability of HF services** |  |  |  |  |
| Yes | 355 (72.3) | 136 (27.7) | Reference |  |
| No | 30 (57.7) | 22 (42.3) | 1.9 (1.07 – 3.44) | 0.029 |
| **Attitude of healthcare providers** |  |  |  |  |
| Positive | 355 (74.0) | 125 (26.0) | Reference |  |
| Negative | 15 (44.1) | 19 (55.9) | 3.5 (1.77 – 7.30) | < .001 |
| Neutral | 15 (51.7) | 14 (48.3) | 2.7 (1.24 – 5.65) | 0.012 |
| **Received counselling** |  |  |  |  |
| Yes | 329(80.8) | 78(19.2) | Reference |  |
| No | 56 (41.2) | 80 (58.8) | 6 (3.95 – 9.18) | < .001 |
| **Felt judged by while seeking care because of age** |  |  |  |  |
| No | 332 (80.8) | 79 (19.2) | Reference |  |
| Yes | 53 (40.2) | 79 (59.8) | 6.2(4.09 – 9.59) | < .001 |
| **Are there costs involved in accessing healthcare services** |  |  |  |  |
| No | 245 (77.8) | 70 (22.2) | Reference |  |
| Yes | 140 (61.4) | 88 (38.6) | 2.2 (1.51 – 3.21) | < .001 |
| **Community related factors** |  |  |  |  |
| **Place of Residence** |  |  |  |  |
| Urban | 165 (78.2) | 46 (21.8) | Reference |  |
| Rural | 220 (66.3) | 112(33.7) | 1.8 (1.23 - 2.72) | 0.003 |
| **Do media talk about issues related to teenage pregnancy** |  |  |  |  |
| No | 54 (73.0) | 20 (27.0) | Reference |  |
| Yes | 331 (70.6) | 138 (29.4) | 1.1 (0.65 – 1.95) | 0.673 |
| **Cultural influence including early marriage practices** |  |  |  |  |
| No | 227 (68.6) | 104 (31.4) | Reference |  |
| Yes | 158 (74.5) | 54 (25.5) | 0.7 (0.51 – 1.10) | 0.137 |
| **Religious influence including early marriage practices** |  |  |  |  |
| No | 118 (68.2) | 55 (31.8) | Reference |  |
| Yes | 267 (72.2) | 103 (27.8) | 0.8 (0.56 – 1.23) | 0.345 |
| **Peer influence including early marriage practices** |  |  |  |  |
| Yes | 142 (62.6) | 85 (37.4) | Reference |  |
| No | 243 (76.9) | 73 (23.1) | 0.5 (0.34 - 73) | < .001 |

\*significant at $P$ value $<0.05$ , Abbreviations: OR, odds ratio; CI, confidence interval.

### Family related factors

All family related factors were significantly associated with teenage pregnancy at a bivariate level of analysis. Teenagers neglected by parents exhibited a significantly high prevalence of teenage pregnancies 60.0% and were 7.7 times more likely to get pregnant than their counterparts who did not experience parental neglect (OR 7.7, 95%CI 5.12-11.79). Similarly, other family related factors associated with teenage pregnancy were; do not discuss SRH with parents, parents talk about sex education, witnessed domestic violence, do not stays with parents, and low socioeconomic status of family (Table 3).

### Health facility factors

At a bivariate level of analysis, all health facility related factors were significantly associated with teenage pregnancy. There was a significantly high prevalence of teenage pregnancy 59.8% among teenagers who felt judged by healthcare service providers/healthcare workers while seeking care because of their age compared to 19.2% among those who did not feel judged. Teenagers who felt judged by healthcare workers while seeking care because of their age were 6.2 times more likely to get pregnant compared to those who did not feel judged (OR 6.2, 95%CI 4.09-9.59). Similarly, other teenage pregnancy associated health facility factors included; no provision of youth tailored health services, affordability of HF services, attitude of healthcare providers, not receiving counselling, costs involved in accessing HCS (Table 3).

### Community related factors

Place of residence and peer influence including early marriage practices were the only community related factors significantly associated with teenage pregnancy at a bivariate level of analysis. Teenage pregnancy was more prevalent among teenagers in rural areas (33.7%) than their counter urban settings (21.8%) with those in rural areas being more likely to get pregnant 1.8 times higher those in urban areas (OR 1.8, 95%CI 1.23-2.72). Community related factors like Religious influence including early marriage practices, Cultural influence including early marriage practices, media help mitigate issue of teenage pregnancy, and, media talk about teenage pregnancy were insignificantly associated with teenage pregnancy in Hoima district (Table 3).

### Multivariable analysis

Table 4 presents the results of multivariable analysis of factors associated with teenage pregnancy. All variables with P values <0.05 at the bivariate level of analysis were selected for the multivariable analysis. The final model depicts the best fit for this analysis. All variables that remained in the multivariable model were statistically significant and they’re thus significantly associated with teenage pregnancy among teenagers in Hoima district.

**Table 4.**
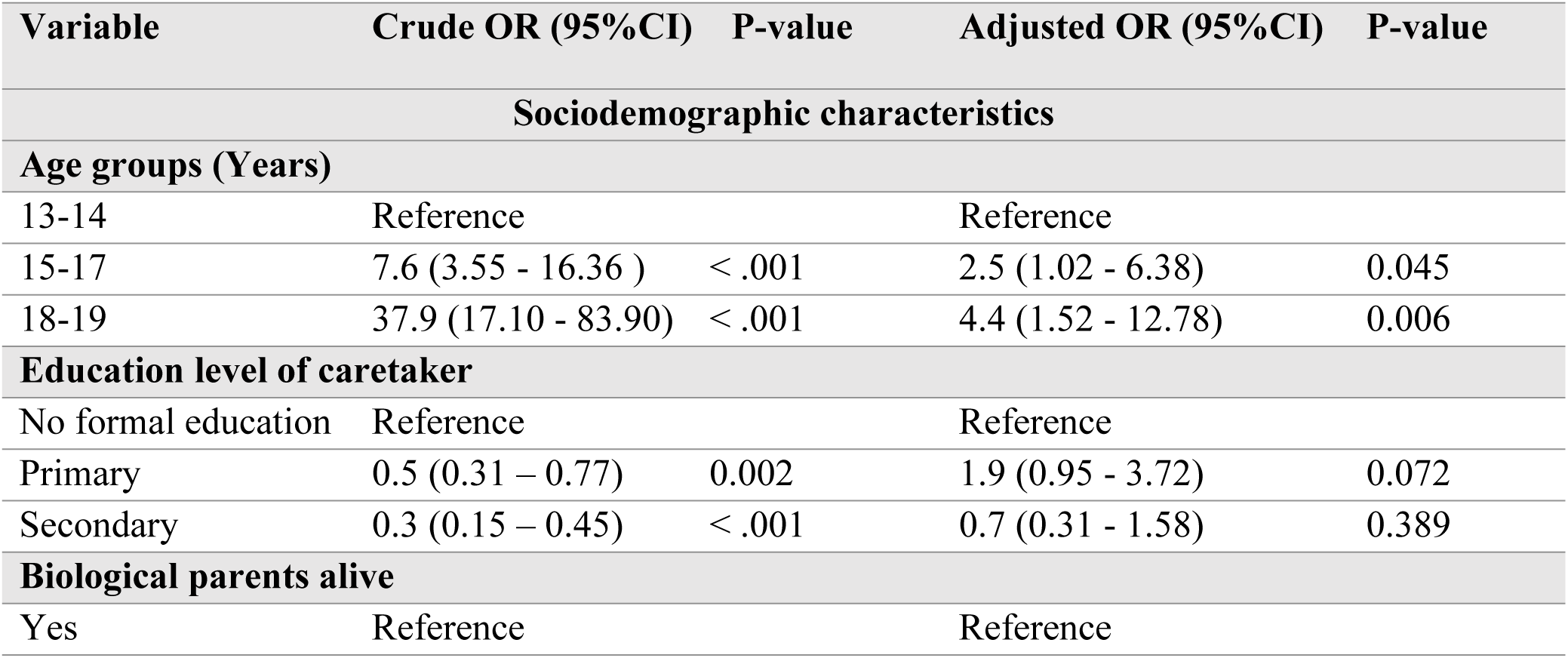
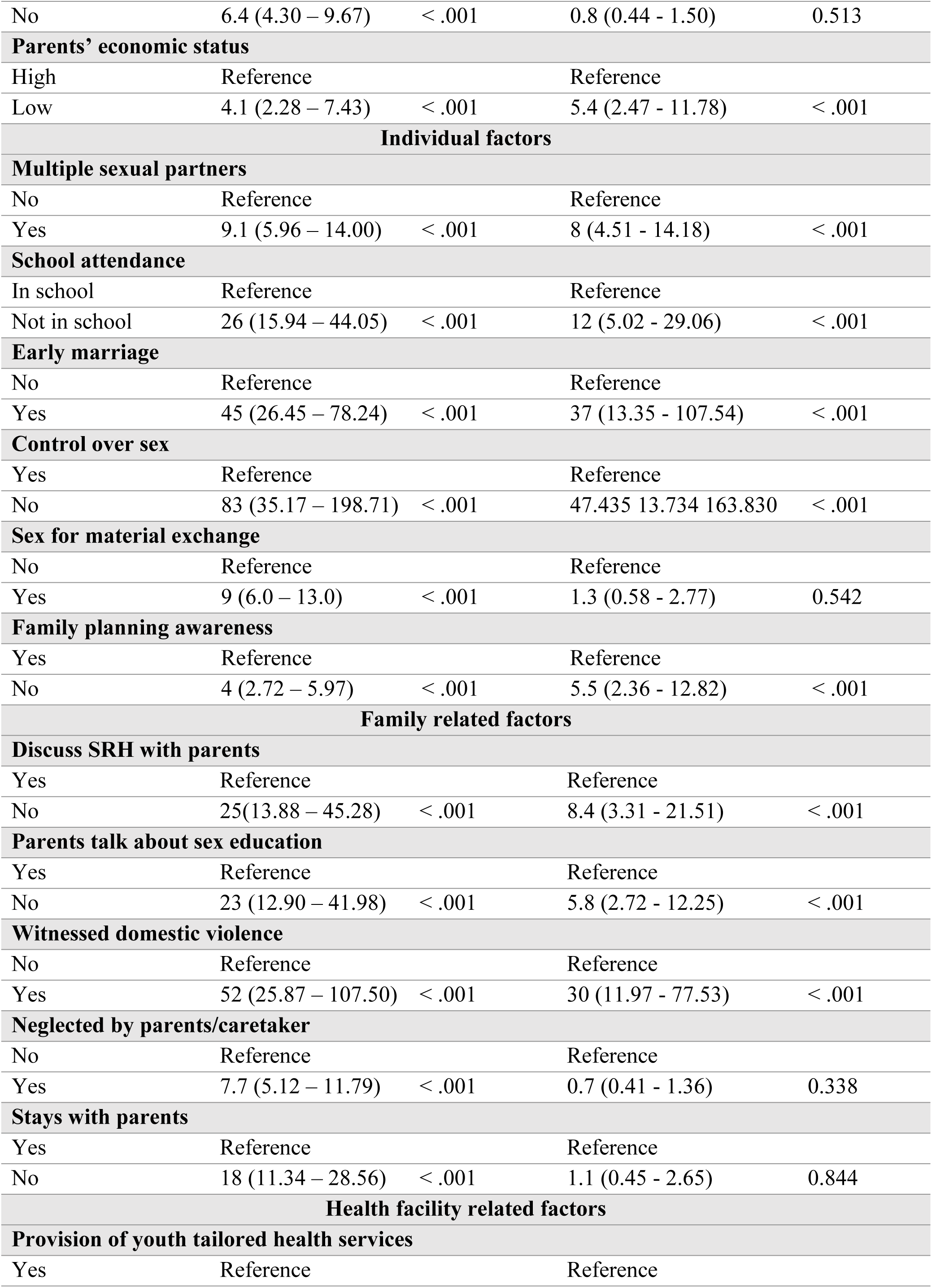
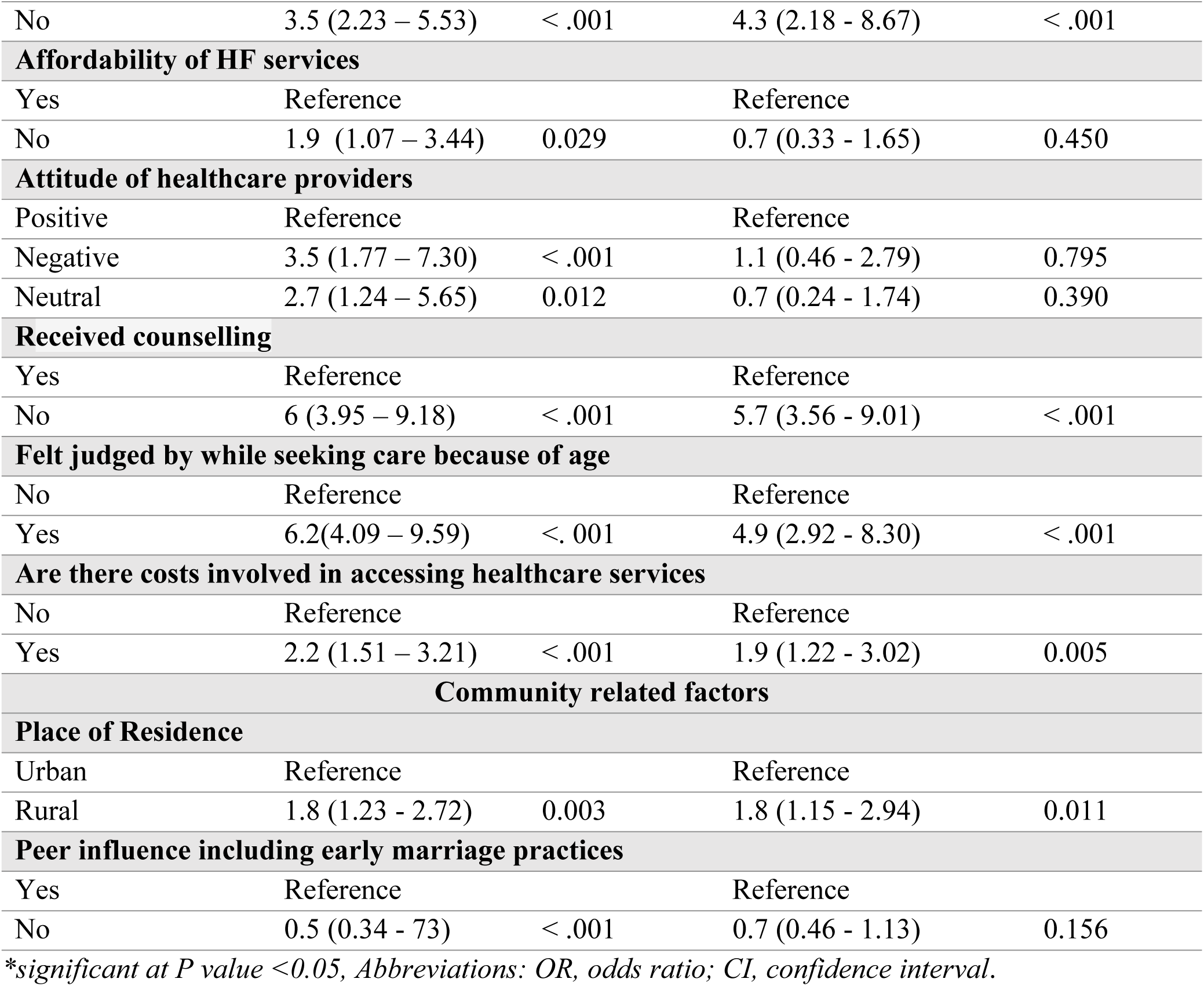
Multivariable analysis of factors associated with teenage pregnancy.

| Variable | Crude OR (95%CI) | P-value | Adjusted OR (95%CI) | P-value |
| --- | --- | --- | --- | --- |
| **Sociodemographic characteristics** |  |  |  |  |
| **Age groups (Years)** |  |  |  |  |
| 13-14 | Reference |  | Reference |  |
| 15-17 | 7.6 (3.55 - 16.36 ) | < .001 | 2.5 (1.02 - 6.38) | 0.045 |
| 18-19 | 37.9 (17.10 - 83.90) | < .001 | 4.4 (1.52 - 12.78) | 0.006 |
| **Education level of caretaker** |  |  |  |  |
| No formal education | Reference |  | Reference |  |
| Primary | 0.5 (0.31 – 0.77) | 0.002 | 1.9 (0.95 - 3.72) | 0.072 |
| Secondary | 0.3 (0.15 – 0.45) | < .001 | 0.7 (0.31 - 1.58) | 0.389 |
| **Biological parents alive** |  |  |  |  |
| Yes | Reference |  | Reference |  |
| No | 6.4 (4.30 – 9.67) | < .001 | 0.8 (0.44 - 1.50) | 0.513 |
| **Parents' economic status** |  |  |  |  |
| High | Reference |  | Reference |  |
| Low | 4.1 (2.28 – 7.43) | < .001 | 5.4 (2.47 - 11.78) | < .001 |
| **Individual factors** |  |  |  |  |
| **Multiple sexual partners** |  |  |  |  |
| No | Reference |  | Reference |  |
| Yes | 9.1 (5.96 – 14.00) | < .001 | 8 (4.51 - 14.18) | < .001 |
| **School attendance** |  |  |  |  |
| In school | Reference |  | Reference |  |
| Not in school | 26 (15.94 – 44.05) | < .001 | 12 (5.02 - 29.06) | < .001 |
| **Early marriage** |  |  |  |  |
| No | Reference |  | Reference |  |
| Yes | 45 (26.45 – 78.24) | < .001 | 37 (13.35 - 107.54) | < .001 |
| **Control over sex** |  |  |  |  |
| Yes | Reference |  | Reference |  |
| No | 83 (35.17 – 198.71) | < .001 | 47.435 13.734 163.830 | < .001 |
| **Sex for material exchange** |  |  |  |  |
| No | Reference |  | Reference |  |
| Yes | 9 (6.0 – 13.0) | < .001 | 1.3 (0.58 - 2.77) | 0.542 |
| **Family planning awareness** |  |  |  |  |
| Yes | Reference |  | Reference |  |
| No | 4 (2.72 – 5.97) | < .001 | 5.5 (2.36 - 12.82) | < .001 |
| **Family related factors** |  |  |  |  |
| **Discuss SRH with parents** |  |  |  |  |
| Yes | Reference |  | Reference |  |
| No | 25(13.88 – 45.28) | < .001 | 8.4 (3.31 - 21.51) | < .001 |
| **Parents talk about sex education** |  |  |  |  |
| Yes | Reference |  | Reference |  |
| No | 23 (12.90 – 41.98) | < .001 | 5.8 (2.72 - 12.25) | < .001 |
| **Witnessed domestic violence** |  |  |  |  |
| No | Reference |  | Reference |  |
| Yes | 52 (25.87 – 107.50) | < .001 | 30 (11.97 - 77.53) | < .001 |
| **Neglected by parents/caretaker** |  |  |  |  |
| No | Reference |  | Reference |  |
| Yes | 7.7 (5.12 – 11.79) | < .001 | 0.7 (0.41 - 1.36) | 0.338 |
| **Stays with parents** |  |  |  |  |
| Yes | Reference |  | Reference |  |
| No | 18 (11.34 – 28.56) | < .001 | 1.1 (0.45 - 2.65) | 0.844 |
| **Health facility related factors** |  |  |  |  |
| **Provision of youth tailored health services** |  |  |  |  |
| Yes | Reference |  | Reference |  |
| No | 3.5 (2.23 – 5.53) | < .001 | 4.3 (2.18 - 8.67) | < .001 |
| **Affordability of HF services** |  |  |  |  |
| Yes | Reference |  | Reference |  |
| No | 1.9 (1.07 – 3.44) | 0.029 | 0.7 (0.33 - 1.65) | 0.450 |
| **Attitude of healthcare providers** |  |  |  |  |
| Positive | Reference |  | Reference |  |
| Negative | 3.5 (1.77 – 7.30) | < .001 | 1.1 (0.46 - 2.79) | 0.795 |
| Neutral | 2.7 (1.24 – 5.65) | 0.012 | 0.7 (0.24 - 1.74) | 0.390 |
| **Received counselling** |  |  |  |  |
| Yes | Reference |  | Reference |  |
| No | 6 (3.95 – 9.18) | < .001 | 5.7 (3.56 - 9.01) | < .001 |
| **Felt judged by while seeking care because of age** |  |  |  |  |
| No | Reference |  | Reference |  |
| Yes | 6.2(4.09 – 9.59) | < .001 | 4.9 (2.92 - 8.30) | < .001 |
| **Are there costs involved in accessing healthcare services** |  |  |  |  |
| No | Reference |  | Reference |  |
| Yes | 2.2 (1.51 – 3.21) | < .001 | 1.9 (1.22 - 3.02) | 0.005 |
| **Community related factors** |  |  |  |  |
| **Place of Residence** |  |  |  |  |
| Urban | Reference |  | Reference |  |
| Rural | 1.8 (1.23 - 2.72) | 0.003 | 1.8 (1.15 - 2.94) | 0.011 |
| **Peer influence including early marriage practices** |  |  |  |  |
| Yes | Reference |  | Reference |  |
| No | 0.5 (0.34 - 73) | < .001 | 0.7 (0.46 - 1.13) | 0.156 |

\*significant at $P$ value $<0.05$ , Abbreviations: OR, odds ratio; CI, confidence interval.

### Factors associated with teenage pregnancy

The factors that remained significantly associated with increased odds of teenage pregnancy at multivariate analysis level included; age (18-19 years) (AOR 4.4, 95%CI 1.52–12.78, p = 0.006), low parents’ economic status (AOR 5.4, 95%CI 2.47–11.78, p< 0.001), multiple sexual partners (AOR 8, 95%CI 4.51–14.18, p< 0.001), being out of school (AOR 12, 95%CI 5.02–29.06, p< 0.001), early marriage (AOR 37, 95%CI 13.35–107.54, p<0.001), having no control over sex (AOR 47, 95%CI 13.7-163.8, p< 0.001), not discussing SRH with parents (AOR 8.4, 95%CI 3.31–21.51, p< 0.001), witnessed domestic violence (AOR 30, 95%CI 11.97–77.53, p< 0.001), never received counselling (AOR 5.7, 95%CI 3.56–9.01, p = 0.001), and, rural residence (AOR 1.8, 95%CI 1.15–2.94, p = 0.011) (Table 4).

Caretakers’ level of education (Secondary: AOR 0.7, 95%CI 0.31 – 1.58, P-value = 0.389), and no peer influence including early marriage practices (AOR 0.7, 95%CI 0.46 – 1.13, P-value 0.156) were associated with decreased odds of teenage pregnancy (Table 4).

Affordability of HF services, attitude of healthcare providers, peer influence including early marriage practices, stays with parents, neglected by parents, education level of caretaker, biological parents alive, and, sex for material exchange factors were insignificantly associated with teenage pregnancy (Table 4).

## Discussion

This study determined the prevalence of teenage pregnancies and described the demographic, individual, family, community and Health facility factors associated with pregnancy among teenage girls aged 13–19 years in Hoima district, Uganda. In this study, the estimated prevalence of teenage pregnancy was 29%, above 2.8% from a similar study in Agago district [13], national average of 24% and slightly below 30.6% reported in a similar study [12]. The observed difference in the teenage pregnancy prevalence could have been due to differences in the measurement of outcome. In this study, teenage pregnancy was based on self-report which is unreliable and may under or overestimate the true burden of teenage pregnancies while in a study by Okot [13], teenage pregnancy was determined by rapid hCG test for urine. These differences can as well be explained by the methodological approaches used, the Demographic Health Survey (DHS) report included a larger sample size and it was done over five years before the current study which used a cross section study design with considerable less sample size compared to national survey.

## Demographic factors

The prevalence of teenage pregnancy increased with an increase in age. Accordingly, the prevalence of teenage pregnancy among aged 13–14 years old was 4.7% compared to those between 18 and 19 years old (64.9%). Similarly, 77.1% of teenagers 18–19 years were married compared to 1.2% of the teenagers aged 13–14 years old. This may be explained as age increases the probability of sexual intercourse and marriage also increases; as a result, the risk of exposure to pregnancy and childbearing also increases. This finding is consistent with study conducted in Ethiopia which indicated that as the age of the adolescent increased, the odds of an adolescent pregnancy also increased [15]. In this study, older age groups (15-17 years and 18-19 years) had significantly higher odds of teenage pregnancy compared to the 13-14 year age group (AOR 2.5, 95% CI 1.02-6.38 and AOR 4.4, 95% CI 1.52-12.78, respectively). This finding is consistent with previous studies that have demonstrated an increased risk of teenage pregnancy with advancing age [15].

This study found that parents’ economic status played a significant influence on teenage pregnancy as teenagers whose parents were poor were 5.4 times (AOR 5.4, 95% CI 2.47-11.78) more likely to get pregnancy than those whose parents had good economic status. This finding is in corroboration with the well-established link between poverty and increased vulnerability to teenage pregnancy [16, 17]. A similar study in Uganda also reported similar findings where teenagers from families with a low income had a significant risk of early sexual activity involvement and teen pregnancy [11]. A study in Iran reported that the majority of teenagers from families with poor economic background find it difficult to meet their expectations posing the risk of early involvement in sexual activity which not only cause early pregnancy but also increase the risk of sexually transmitted infections and other life-threatening maternal complications and mortality [18]. This can however be explained from different angles, low income is linked with failure to obtain basic needs including clothes, food, and entertainment which creates an environment for dropping out of school, some run away from homes looking for jobs, increase risk of raping and some get easy to accept men hoping for support that all do creates an atmosphere for early sexual practices and pregnancy [17].

## Individual factors

Not attending school was associated with substantially higher odds of teenage pregnancy (AOR 12.0, 95% CI 5.02-29.06). This finding aligns with similar studies in Ethiopia and Sri Lanka which showed that being out of school was significantly associated with pregnancy [14, 19]. Formal education equip teenagers with crucial knowledge through comprehensive sexual education hence empowering them to negotiate their sexuality, understand contraception, and make informed choices about childbearing [20]. Staying in school offers a benefit for girls in reducing teen pregnancy by providing a structured environment with supervision which potentially reduces opportunities for early sexual activity [15]. Furthermore, education strengthens a girl’s stance in deciding about her reproductive future, such as the timing and number of children she desires. Compared to their educated counterparts, girls without schooling are less able to negotiate sex with partners, potentially leading to earlier marriage and increased risk of gender-based violence [15, 19].

Despite the legislative measures to prevent early marriage, the practice persists in Uganda and several SSA countries [20]. This study showed early marriage was associated with teenage pregnancy which concurs with a study in Uganda [21]. During early marriage, teenagers are exposed to frequent and unprotected sexual activity which often leads to an early and risky first birth. Gideon [21], discussed early marriage as a precursor for teenage pregnancy owing to the strong expectations that fertility was proved by conceiving within a year of marriage.

There was a strong link between teenage pregnancies and having multiple sexual partners among teenage girls and this was supported by the fact that teenage girls with multiple sexual partners were 8 times (AOR 8.0, 95% CI 4.51-14.18) more likely to get pregnant compared to their counterparts with no multiple sexual partners. This finding was in agreement with findings in study which also reported that risky sexual behaviors like having multiple sexual partners increased the odds of teenage girls getting unwanted pregnancies [14]. This can however be explained from different angles; Firstly, having multiple sexual partners may lead to more frequent sexual activity, which even with contraceptive use, heightens the overall chances of pregnancy due to the possibility of inconsistent or incorrect use [22]. Secondly, contraceptive methods are known not to be 100% effective, and with multiple partners, the compounded risk of contraceptive failure like condom breakage or missed pills multiplies the chances of unintended pregnancy [14, 22].

Failure to have control over sexual decisions was a significant predictor of teenage pregnancy in this study where teenage girls who did have control over sexual decision were 47 times (AOR 47, 95% CI 13.73-163.83) more likely to report teenage pregnancy than their counterparts who had control over sexual decision. This finding was consistent with the finding by Barnert who reported heightened pregnancy prevalence among women who had little say in matters related to sex [23]. Lack of control from the teenage girls over sex can lead to coercion and unwanted sexual encounters hence non-consensual and unprotected sexual activity resulting in pregnancy [23]. Furthermore, teenage girls’ access to sexual and reproductive health information and contraceptive methods may also be restricted by financial constraints and manipulation by partners who use deception to engage in unprotected sex [23]. Therefore, the cumulative impact of these factors (coercion, inability to ensure contraceptive use, limited access to services, and potential partner manipulation) arising from this lack of control over sexual decisions severely increases girls’ vulnerability to unintended teenage pregnancy

## Family related factors

Parent-child open communication about sex and reproductive health in this study was significantly associated teenage pregnancy where by teenagers who did not have parent-child communication regarding sex and reproductive health were 8.4 times (AOR 8.4, 95%CI 3.31-21.51) more likely to report teenage pregnancy. In 2020, studies from Zambia and Kenya respectively found that open parent-child communication offered a protective effect against teenage pregnancy which correlates with this study [24, 25]. Lack of open parent-child communication leaves teenage girls without reliable information from parents hence relying on inaccurate sources like peers or media, lacking crucial knowledge about safe sexual practices and proper contraceptive which eventually exposes teenagers to several vulnerabilities that heighten their risk of unintended pregnancy [24].

Witnessing domestic violence was strongly associated with increased odds of teenage pregnancy which is consistent with previous studies that have highlighted the detrimental impact of violence on adolescent well-being and sexual and reproductive health outcomes [26]. Emotional and psychological distress caused by domestic violence can make adolescents more prone to risky sexual behavior and unintended pregnancies.

In this study, witnessing domestic violence was strongly associated with increased odds of teenage pregnancy as teenage who witnessed domestic violence were 30 times (AOR 30, 95% CI 11.97-77.53), more likely to report teenage pregnancy compared to their counterparts who did not witness or experience domestic violence. This finding was in agreement with findings from the previous studies that reported increased occurrence of teenage pregnancy among teenagers who experienced domestic violence at any single point in their life time [26]. This can be explained by the fact that domestic violence causes profound psychological, emotional, and social consequences on teenagers leading to mental health issues like depression, anxiety, and low self-esteem, increasing the likelihood of risky sexual behavior and poor decision-making [26].

## Health facility related factors

With regards to counseling, the results show that teenagers who did not receive counseling where 5.7 times more likely to get pregnant compared to their peers who received counseling while seeking health services at health facilities (AOR 5.7, 95% CI 3.56-9.01). This finding correlates with finding from an earlier study in Nigeria where teenage pregnancy was more prevalent among teenagers with no counselling about sexual and reproductive health. Through counselling, teenagers get accurate information, advice, and support regarding sexual and reproductive health services that enable teenage girls to make informed sexual and reproductive health decisions and consequently reduce her chances of teenage pregnancy [27].

Having costs associated with accessing healthcare services was a significant risk factor for teenage pregnancy (AOR 1.9, 95% CI 1.22-3.02), which concurs with studies that have emphasized the importance of affordable and accessible sexual and reproductive health services for young people for adolescents [28]. Financial barriers can limit adolescents’ ability to access essential healthcare services, including contraceptive counseling and provision, increasing their risk of unintended pregnancy [28]. High costs attached to healthcare services could discourage teenagers, especially those from a poor economic background from seeking professional guidance about sexual health. These associated costs of transportation, combined with potential lost income due to missed work or school, can be a significant barrier [16].

In addition, being looked down upon by healthcare workers due to age while seeking care was significantly associated with high odds of teenage pregnancy (AOR 4.9, 95% CI 2.92-8.30). Teenagers who felt judged were 4.9 times more likely to get pregnant than their counterparts. This can be linked to the fact that the perceived judgement by the teenagers creates a barrier to accessing essential SRHs; breaking trust and confidentiality, leading to teenagers withholding information; and causing detrimental psychological effects hence affecting self-esteem, decision-making abilities, and vulnerability to risky sexual behavior increasing the chances of getting pregnant. This study also showed a significant association between teenage pregnancy and the provision of youth tailored health services at health facilities. Lack of youth-tailored health services was associated with higher odds of teenage pregnancy (AOR=4.3, CI=2.18-8.67), which is in line with other studies emphasizing the importance of adolescent-friendly SRH services [11, 21].

## Community related factor

This study showed a significantly high prevalence of teenage pregnancy among teenage girls from rural areas than those from urban settings. Teenage girls from rural areas where 1.8 times (AOR 1.8, 95%CI 1.15 – 2.94, p-value 0.011) more likely to get pregnant than their counterparts in urban areas. In 2020, a similar study by in Zambia reported that teenage pregnancies were more common among teenage girls residing in rural areas than urban areas which is consistent with the present study [25]. This is because people living in rural settings may not have sufficient access to healthcare facilities. Another reason could be that girls from less developed communities and rural areas are least likely to access quality education as well as sound sexual and reproductive health services that provide adequate information on birth control. To the contrary, in 2021, a multi-country analysis study on teenage pregnancy reported lower odds of having first pregnancy among teenager from West African sub-region who lived in rural areas [2]. This could be because teenage girls from rural areas are less exposed to pornographic content which could expose them to risky sexual behaviors.

## Conclusions

This study explored the factors associated with teenage pregnancy in Hoima district, Uganda. The teenage pregnancy estimate of 29% in this study is higher than the national average of 24%. Teenager’s age, type of residence, parents’ economic status, domestic violence, provision of youth tailored health services, school attendance, early marriage, sex for material exchange, and, multiple sexual partners were the most important risk factors of teenage pregnancy among teenage girls in Hoima district, Uganda.

## Declarations

### Ethics approval and consent to participate

Ethical approval was sought from the Research and Ethics committee of Makerere University School of public Health. The study was conducted in accordance with the ethical principles of the Declaration of Helsinki. Permission to conduct the study was sought from the Chief Administrative Officer (CAO), Resident District commissioner (RDC) and the office of Local councils, Hoima district. The purpose of the study was explained and information obtained was used only for research purposes. Written informed consent was obtained from study participants before conducting any interviews and confidentiality was ensured throughout the study. Permission was also sought from public health department, Hoima district and village leaders in the selected villages. Individual study respondents and their parents were approached to obtain their consent to carry out the study. All information provided by the respondents was confidential.

## Availability of data and materials

The data set supporting the findings is available upon request from the corresponding author

## Consent for publication

Not applicable.

## Author contributions

W.J.W and R.K conceived the study, designed the research, collected the data, and drafted the manuscript. S.K, J.B and, R.K supervised, provided critical revisions, and provided expert guidance throughout the study design, data interpretation, and manuscript preparation. R.K analyzed the data, developed the visualizations, and wrote the results section of the manuscript. All authors read and approved the final manuscript for submission.

## Data Availability

The data set supporting the findings is available upon request from the corresponding author

## List of abbreviations and acronyms

DHS: Demographic Health Survey
SSA: Sub-Saharan Africa
UBOS: Uganda Bureau of Statistics
UNICEF: United Nations International Children’s Emergency Fund
WHO: World Health Organization
PRB: Population Reference Bureau
SRH: Sexual and Reproductive Health
OR: Odds Ratio
AOR: Adjusted Odds Ratio

## Acknowledgements

We acknowledge the Hoima district administrative authorities and the District Health Team (DHT) for permitting us to conduct the study.

## References

1. United Nations Children’s Fund, UNICEF: early childbearing and teenage pregnancy rates by country; 2023. https://data.unicef.org/topic/child-health/adolescent-health/. Accessed November 12, 2024.

2. Ahinkorah BO, Kang M, Perry L, Brooks F, Hayen A. Prevalence of first adolescent pregnancy and its associated factors in sub-Saharan Africa: A multi-country analysis. PloS one. 2021 Feb 4;16(2):e0246308.

3. WHO 2022. Early marriages, adolescent and young pregnancies. Geneva: WHO, 1–4.

4. Asare BY, Baafi D, Dwumfour-Asare B, Adam AR. Factors associated with adolescent pregnancy in the Sunyani Municipality of Ghana. International Journal of Africa Nursing Sciences. 2019 Jan 1;10:87–91.

5. UBOS 2022. https://www.ubos.org/uganda-demographic-and-health-survey-udhs-2022-key-findings/

6. Atuyambe LM, Kibira SP, Bukenya J, Muhumuza C, Apolot RR, Mulogo E. Understanding sexual and reproductive health needs of adolescents: evidence from a formative evaluation in Wakiso district, Uganda. Reproductive health. 2015 Apr 22;12(1):35.

7. Manzi F, Ogwang J, Akankwatsa A, Wokali OC, Obba F, Bumba A, Nekaka R, Gavamukulya Y. Factors associated with teenage pregnancy and its effects in Kibuku Town Council, Kibuku District, Eastern Uganda: A cross sectional study. Primary Health Care Open Access. 2018;8(02).

8. PRB 2017. Population Reference Bureau. World Population Data sheets with special focus on the youth (2017).

9. Kassa GM, Arowojolu AO, Odukogbe AA, Yalew AW. Prevalence and determinants of adolescent pregnancy in Africa: a systematic review and meta-analysis. Reproductive health. 2018 Nov 29;15(1):195.

10. WHO 2023. Adolescent pregnancy fact sheet. Adolescent Pregnancy Fact Sheet, 1–4.

11. Ochen AM, Chi PC, Lawoko S. Predictors of teenage pregnancy among girls aged 13– 19 years in Uganda: a community based case-control study. BMC pregnancy and childbirth. 2019 Jun 24;19(1):211.

12. Musinguzi M, Kumakech E, Auma AG, Akello RA, Kigongo E, Tumwesigye R, Opio B, Kabunga A, Omech B. Prevalence and correlates of teenage pregnancy among in-school teenagers during the COVID-19 pandemic in Hoima district western Uganda–A cross sectional community-based study. PloS one. 2022 Dec 16;17(12):e0278772.

13. Okot C, Laker F, Apio PO, Madraa G, Kibone W, Pebalo Pebolo F, Bongomin F. Prevalence of teenage pregnancy and associated factors in Agago district, Uganda: a community-based survey. Adolescent health, medicine and therapeutics. 2023 Dec 31:115–24.

14. Mezmur H, Assefa N, Alemayehu T. Teenage pregnancy and its associated factors in eastern Ethiopia: a community-based study. International Journal of Women’s Health. 2021 Feb 26:267–78.

15. Ayele BG, Gebregzabher TG, Hailu TT, Assefa BA. Determinants of teenage pregnancy in Degua Tembien District, Tigray, Northern Ethiopia: A community-based case-control study. PloS one. 2018 Jul 25;13(7):e0200898.

16. Kooko R, Wafula ST, Orishaba P. Socioeconomic determinants of malaria prevalence among under five children in Uganda: Evidence from 2018-19 Uganda Malaria Indicator Survey. Journal of Vector Borne Diseases. 2023 Jan 1;60(1):38–48.

17. Moshi FV, Tilisho O. The magnitude of teenage pregnancy and its associated factors among teenagers in Dodoma Tanzania: a community-based analytical cross-sectional study. Reproductive Health. 2023 Feb 3;20(1):28.

18. Montazeri S, Gharacheh M, Mohammadi N, Alaghband Rad J, Eftekhar Ardabili H. Determinants of early marriage from married girls’ perspectives in Iranian setting: a qualitative study. Journal of environmental and public health. 2016;2016(1):8615929.

19. Dulitha F, Nalika G, Upul S, Chrishantha WM, De Alwis SR, Hemantha S, Chithramalee DS. Risk factors for teenage pregnancies in Sri Lanka: perspective of a community based study. Health Science Journal. 2013 Jul 1;7(3):269.

20. Yaya S, Odusina EK, Bishwajit G. Prevalence of child marriage and its impact on fertility outcomes in 34 sub-Saharan African countries. BMC international health and human rights. 2019 Dec 19;19(1):33.

21. Gideon R. Factors associated with adolescent pregnancy and fertility in Uganda: analysis of the 2011 demographic and health survey data. American Journal of Sociological Research. 2013;3(2):30–5.

22. Trussell, J., Raymond, E.G. and Cleland, K., 2014. Emergency Contraception: A Last Chance to Prevent Unintended Pregnancy. Contemporary Readings in Law & Social Justice, 6(2).

23. Barnert ES, Godoy SM, Hammond I, Kelly MA, Thompson LR, Mondal S, Bath EP. Pregnancy outcomes among girls impacted by commercial sexual exploitation. Academic pediatrics. 2020 May 1;20(4):455–9.

24. Isaksen KJ, Musonda P, Sandøy IF. Parent-child communication about sexual issues in Zambia: a cross sectional study of adolescent girls and their parents. BMC public health. 2020 Jul 16;20(1):1120.

25. Maina BW, Ushie BA, Kabiru CW. Parent-child sexual and reproductive health communication among very young adolescents in Korogocho informal settlement in Nairobi, Kenya. Reproductive health. 2020 Jun 1;17(1):79.

26. Miura PO, Tardivo LS, Barrientos DM, Egry EY, Macedo CM. Adolescence, pregnancy and domestic violence: social conditions and life projects. Revista Brasileira de Enfermagem. 2020 Jun 1;73(Suppl 1):e20190111.

27. Akanbi MA, Ope BW, Adeloye DO, Amoo EO, Iruonagbe TC, Omojola O. Influence of socio-economic factors on prevalence of teenage pregnancy in Nigeria. African Journal of Reproductive Health. 2021;25(5s):138–46.

28. Guttmacher Institute. (2019, April). How much does it cost to get contraception? https://www.guttmacher.org/report/adding-it-up-investing-in-sexual-reproductive-health-2019

